# Cardiac amyloidosis and the risk of Alzheimer’s disease: a population-based nested case-control study

**DOI:** 10.64898/2026.01.16.26344270

**Authors:** Fannie Bretelle, Frederic Gervais, Virginie Dauphinot, Michel Chuzeville, Isabelle Quadrio, Virginie Desestret, Antoine Garnier-Crussard

**Affiliations:** Clinical Research Center Ageing-Brain-Frailty, Lyon Institute For Aging, Charpennes Hospital, Hospices Civils de Lyon, 69100 Villeurbanne, France; Geriatric Cardiology Department, Edouard Herriot Hospital, Hospices Civils de Lyon, Lyon, France; Department of Pharmacy, Charpennes Hospital, Hospices Civils de Lyon, Villeurbanne, France; Clinical and Research Memory Centre of Lyon, Lyon Institute For Aging, Hospices Civils de Lyon, Lyon 1 University, 69100 Villeurbanne, France; Biochemistry and Molecular Biology Department Neurodegenerative Pathologies LBMMS Hospices Civils de Lyon LyonFrance; INSERM U1314/UMR CNRS5284, SynatAc Team, MeLis Institute, Lyon, France; IFrench Reference Center on Paraneoplastic Neurological Syndromes, Hospices Civils de Lyon, Lyon, France; University of Lyon, Université Claude-Bernard Lyon 1, Lyon, France; Université Claude Bernard Lyon 1, CNRS, INSERM, Centre de Recherche en Neurosciences de Lyon CRNL U1028 UMR5292, F-69500, Bron, France

**Author notes:** <u>Corresponding author</u>: Dr. Antoine Garnier-Crussard, Address: Hôpital des Charpennes, 27 rue Gabriel Péri, 69100 Villeurbanne, France. co-first author (Fannie Bretelle, Frederic Gervais).

**Keywords:** Alzheimer’s disease, Transthyretin amyloidosis, Amyloidosis, Dementia, Epidemiology

## Abstract

**Background:** Transthyretin amyloidosis cardiomyopathy (ATTR-CM) and Alzheimer’s disease (AD) are age-related disorders characterized by pathological protein aggregation. Despite shared risk factors and mechanisms, the relationship between ATTR-CM and AD remains poorly understood.

**Methods:** We performed a population-based case–control study using the French National Health Data System from 2019 to 2023. Individuals aged 65 years or older diagnosed with ATTR-CM were matched with controls without ATTR-CM by age, sex, hypertension status, and area of residence. The main exposure was a diagnosis of AD within five years preceding the index date (ATTR-CM diagnosis or equivalent for controls). Conditional logistic regression estimated adjusted odds ratios (ORs) for the association between ATTR-CM and AD, accounting for major dementia risk factors including cardiovascular, metabolic, psychiatric, and lifestyle variables.

**Results:** Among 96,200 participants (19,240 ATTR-CM cases and 76,960 controls), 28,990 (30.6%) were women, and the mean (SD) age was 82.3 (6.6) years. The adjusted OR for AD among ATTR-CM patients was 0.65 (99% CI, 0.56–0.75), indicating a lower likelihood of AD compared with controls.

**Conclusions:** This large nationwide study suggests that ATTR-CM is associated with a reduced risk of AD, warranting further investigation into underlying biological mechanisms or possible diagnostic bias.

## Introduction

Transthyretin amyloidosis cardiomyopathy (ATTR-CM) and Alzheimer’s disease (AD) are two age-related disorders characterized by pathological protein aggregation [1], both of which have a substantial impact on quality of life and increase the risk of dependency [2]. Although these diseases are clinically distinct, they share several pathophysiological features, most notably their strong association with aging and the deposition of amyloid material. Both are classified as proteinopathies, resulting from aberrant folding of specific proteins that leads to the formation of insoluble amyloid fibrils, dominated by β-sheet protein aggregates. In AD, extracellular accumulation of β-amyloid peptides and the formation of neurofibrillary tangles composed of hyperphosphorylated tau protein drive progressive synaptic dysfunction and neurodegeneration [3]. In cardiac amyloidosis, myocardial infiltration by amyloid fibrils-originating from either immunoglobulin light chains (in AL amyloidosis) or from transthyretin (in ATTR-CM) in both genetic and wild-type forms, leads to increased myocardial stiffness, resulting in restrictive cardiomyopathy that progresses to congestive heart failure and arrhythmias [4]. The accumulation of amyloid deposits in the extracellular matrix disrupts tissue architecture and impairs organ function, ultimately causing progressive and irreversible loss of function: cognitive decline in AD and heart failure with preserved ejection fraction in ATTR-CM. The rising prevalence of these disorders in older adults underscores the pivotal role of aging in the dysregulation of proteostasis mechanisms, the cellular machinery responsible for maintaining proper protein folding and clearance. This dysregulation of proteostasis mechanisms may be associated in both conditions with several mechanisms, such as inflammation and innate immune activation [5,6] and failure in the clearance of misfolded proteins [7,8].

The putative relationship or potential co-occurrence between these two diseases have not yet been elucidated. On the one hand, chronic heart failure may lead to a higher risk of AD [9] and case report and case series have previously showed co-occurrence of “brain amyloidosis” and “heart amyloidosis” [10],[11],[12], which could lead to the hypothesis of an increased risk of having both conditions, AD and ATTR-CM. On the other hand, some evidence suggests that TTR may exert a protective effect against Alzheimer’s pathology by binding amyloid-beta peptides, which could lead to the hypothesis of an under-risk of having both conditions. However, this does not imply that ATTR-CM is protective, as the biological activities of TTR amyloid and native tetrameric TTR are different, and misfolded and dysfunctional TTR is itself pathogenic [8].

Nevertheless, few studies have specifically investigated the relationship between these two conditions. The present study aims to further elucidate this potential relationship – at the population level - by exploring associations between ATTR-CM and AD in a population-based case-control study using the French national health data system.

## Methods

### Study Design

We conducted a population-based nested case-control study within the electronic healthcare French database (Système National des Données de Santé, SNDS). SNDS is a real-world database which covers 99% of the French population, representing 9% of the European population. This comprehensive data resource contains anonymous data on individuals’ sociodemographic characteristics, healthcare professional visits, drugs dispensation (linked to ATC, Anatomical Therapeutic Chemical), hospital admissions and home hospitalizations, including medical diagnoses and cause of hospitalization (ICD-10, International Classification of Diseases, release 10) and long-term condition (ALD: *Affections de Longue Durée*) also characterized by ICD-10 code.

This study was strictly observational and used fully anonymized data from the French National Health Data System (SNDS), in accordance with the regulations governing non-interventional clinical research in France. All methods were carried out in accordance with relevant guidelines and regulations. Because the study involved neither direct experimentation on humans nor the use of human biological samples, and relied solely on pre-existing anonymized administrative data, neither written informed consent from participants nor authorization from an institutional ethics committee was required under of the French Public Health Code.

The data analyst involved in the present study has permanent authorized access to anonymized SNDS data in accordance with French regulatory procedures. This regulatory framework specifically exempts such studies from institutional review board approval, as no identifiable personal information is accessible and no intervention is performed on human subjects.

### Selection of Cases and Controls

The base cohort included all living patients at the beginning of the study period (January 1, 2019, to December 31,2023).

Case patients were those identified as having a first diagnosis of ATTR-CM during the study period and aged 65 years old and over, excluding patients with a diagnosis of ATTR-CM in 2018. The patient selection for ATTR-CM was realized according to an algorithm recently developed by the French Referral Center for Cardiac Amyloidosis in the SNDS using ICD-10 diagnosis either from at least one hospital stay or associated with ALD status and drugs delivery [13]. Indeed, as there is no specific ICD-10 code for ATTR-CM, the diagnosis of ATTR-CM required both amyloidosis (ICD-10 E85 code or a tafamidis delivery) and a cardiovascular condition (identified by ICD-10 or medical procedure codes related to either heart failure, arrhythmias, conduction disorders or cardiomyopathies), with the exclusion of patients with probable AL or AA amyloidosis. As part of the algorithm, patients with cerebral amyloid angiopathy (CAA) were also excluded in the ATTR-CM group without delivery of tafamidis to ensure that the ICD-10 code E85 is not linked to CAA rather than ATTR-CM. As CAA is strongly associated with AD, we decided to exclude all patients with CAA from our study population to keep the case and control groups comparable and to conduct a sensitivity analysis to assess the impact of this decision. Index date in cases was defined as the date of first dispensation of tafamidis or the date of the first diagnosis of amyloidosis using ICD-10 E85.

Each case patient was matched to 4 controls by age (within 1 year), sex, area of living and an ICD-10 code of hypertension (ICD-10 I10) during the study period or within the 5 years before death. Age, sex and area of living are common matching factors, and hypertension was also added as a major comorbidity in both AD and ATTR-CM. Matching was based on the risk set sampling approach.[14]

### Alzheimer’s Disease

AD status was retrieved in all cases and their matched controls using ICD-10 code F00 (Dementia in Alzheimer disease) and G30 (Alzheimer disease) listed during any hospital stays, home hospitalizations and ALD status within the 5 years before the index date.

### Confounding Variables

We accounted for potential confounding variable identified as risk factors for ATTR-CM or AD. Firstly, as stroke, atrial fibrillation and heart failure are common in ATTR-CM and are associated with an increased risk of dementia, these were added as covariates in the models. Secondly, we accounted for the main dementia risk factors described in the 2024 Lancet standing Commission on dementia prevention, intervention, and care[15]: diabetes, dyslipidemia, obesity, hearing loss, depression, smoking, alcohol use and visual disorders (glaucoma, aged-related macular degeneration codes were included). Traumatic brain injury, physical inactivity, social isolation and air pollution were not accounted because these conditions are not coded. ICD-10 codes from any hospitalizations and ALD status were used to identify these variables during within the 5 years before index date. We also used a history of at least 3 drug dispensations in the 5 years prior to the index date to track diabetes, depression, glaucoma et aged-related macular degeneration using ATC codes (A10, N06A, S01E and S01LA, respectively). As the information on diagnosis using ICD-10 codes is mainly available during a hospital stay in the SNDS database, we also collected the mean number of hospitalizations within the 5 years before the index date to address potential diagnosis bias related to different numbers of hospitalisations in cases and controls groups.

### Statistical Analysis

A conditional logistic regression was conducted to estimate odds ratios (OR) adjusted for the confounding variables. Analyses were performed using SAS, version 9.4 software (SAS Institute, Cary, North Carolina, USA) and R Statistical Software (v4.1.3; R Core Team 2022). All tests were two tailed, and a priori *p* value less than 0.01 was considered to indicate statistical significance.

## Results

### Flow chart of the study population

The definition of cases in this study, based on the French national Health Data System (SNDS) is summarized in **Figure 1**. We initially identified two groups in patients aged 65 years and over: (1) 11,216 individuals with at least one delivery of tafamidis meglumine between 2019 and 2023, and (2) 25,010 patients with a diagnosis of amyloidosis (ICD-10 code E85) but no tafamidis delivery during the same period. In both groups, we retained only patients with a relevant cardiovascular condition (heart failure, arrhythmia, conduction disorder, or cardiomyopathy). This led to the exclusion of 8,726 individuals with no observed cardiovascular disease. We then applied additional exclusion criteria to remove other forms of amyloidosis (n = 6,643), including patients receiving chemotherapy associated with E85, those treated with medications for AL amyloidosis, patients with multiple myeloma, a history of bone marrow transplant, CAA, or a coding pattern suggestive of AA amyloidosis. We also excluded 71 patients with CAA from the tafamidis-treated group. After all exclusions, the final study population included 20,355 prevalent ATTR-CM patients aged 65 years and more between 2019 and 2023. Among them, 1,085 had a diagnosis of ATTR-CM in 2018 and were removed from the final cohort constituted of 19,270 incident ATTR-CM patient aged 65 years and more.

**Figure 1.**
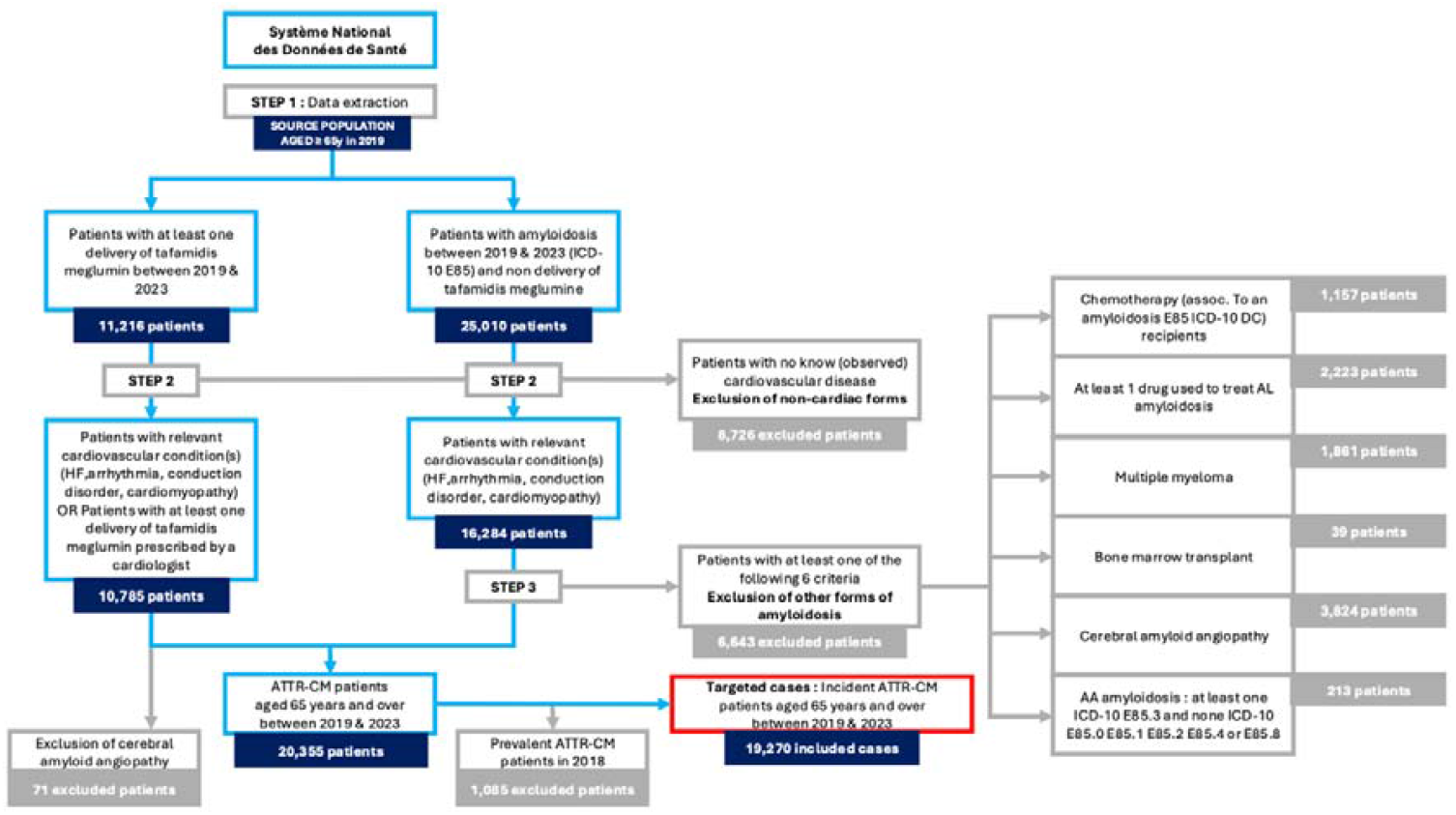
Flow chart of the study population selection from the SNDS (healthcare French database) **Figure legends:** This figure presents the identification process of incident transthyretin cardiac amyloidosis (ATTR-CM) cases aged ≥ 65 years between 2018 and 2023. Step 1: patients aged ≥ 60 years in 2019 were first screened for either tafamidis delivery or a coded diagnosis of amyloidosis between 2019 and 2023. Step 2: two parallel pathways identified (i) patients with at least one tafamidis prescription and (ii) patients with amyloidosis and no tafamidis delivery. In both pathways, individuals were required to have a relevant cardiovascular condition or to initiate tafamidis under cardiologist prescription. At Step 3, prevalent cases were excluded by removing patients with evidence of ATTR-CM before 2018. Additional exclusions were applied for cerebral amyloid angiopathy and for non-ATTR forms of amyloidosis, including chemotherapy-associated amyloidosis, AL amyloidosis, multiple myeloma, bone-marrow transplant, and AA amyloidosis (ICD-10 E85.3, E85.0–E85.5, E85.6, E85.4, E85.8). After applying all exclusion criteria, the final study population included 19 270 incident ATTR-CM cases aged ≥ 65 years between 2018 and 2023.

### Baseline characteristics of cases and controls

Baseline patient characteristics are summarized in **Table 1**. The case group (with ATTR-CM) included 19,240 patients – 30 cases did not match enough controls and were excluded – and the control group included 76,960 patients with a mean (SD) age of 82.3 (6.6) years and 69.4% of males. Atrial fibrillation and heart failure were more prevalent in ATTR-CM patients (69.5% and 74.5%, respectively) compared to controls (24.3% and 22.2%). ATTR-CM patients also had higher rates of stokes, diabetes, dyslipidemia, obesity, alcohol consumption, glaucoma and hearing loss. Patients with depression, age-related macular degeneration, and smoking were similar between cases and controls. There were significantly more hospitalizations in cases than in controls (7.5 and 5.0) in the 5 years before the index date. When comparing AD patients to non-AD patients, the mean number of hospitalizations were similar (**eTable1** in the Supplement).

**Table 1.**
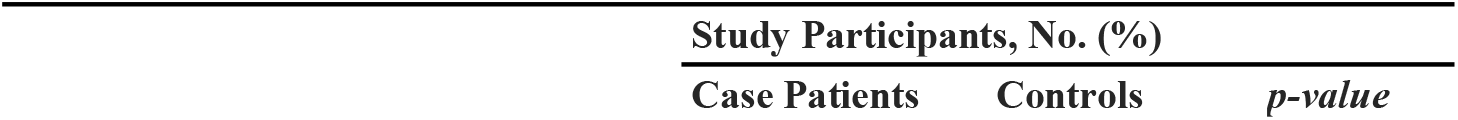
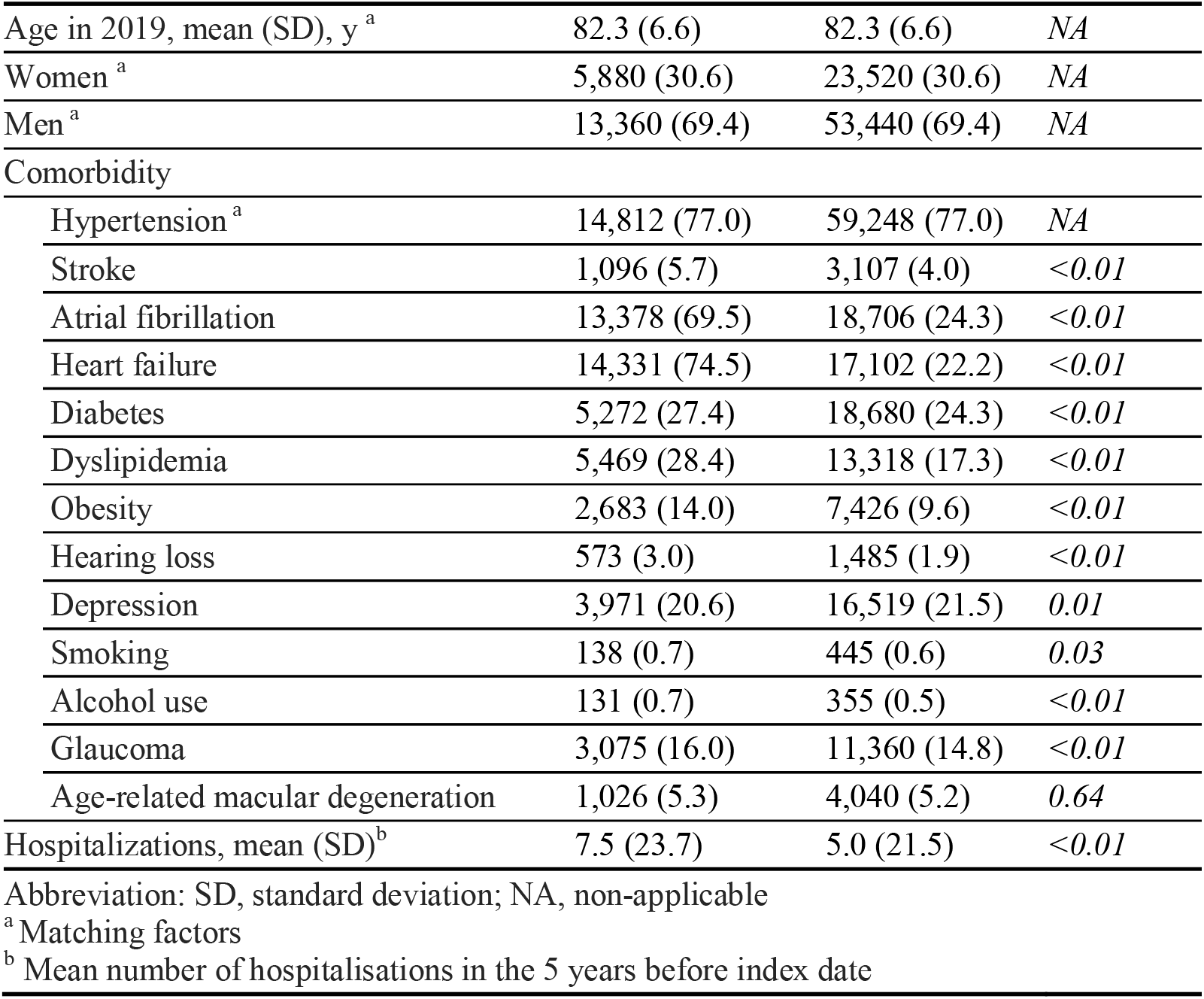
Descriptive Characteristics of Case Patients and Matched Controls.

### Associations between AD and ATTR-CM

The prevalence of AD in the control group was 5.0% and 3.1% in the case group. ATTR-CM was associated with a lower risk of Alzheimer’s disease (adjusted OR 0.65 [99% CI: 0.56– 0.75]). Results are summarized in **Table 2** and full results with adjustments OR associated with confounding variables are available in supplementary files (**eTable2** in the Supplement). Removing all CAA from study population did not change significantly the association in sensitivity analysis (**eTable3** in the Supplement).

**Table 2.** Risk of Alzheimer’s disease associated with cardiac amyloidosis (ATTR-CM) adjusted for covariates.

| Variable | Study participants, No (%) |  | OR (99% CI) |  |
| --- | --- | --- | --- | --- |
|  | Cases (n=19,240) | Controls (n=76,960) | Adjusted | *p-value* |
| Alzheimer's disease |  |  |  |  |
| No | 18,637 (96.9) | 73,127 (95.0) | 1 [Reference] |  |
| Yes | 603 (3.1) | 3,833 (5.0) | 0.65 (0.56-0.75) | *<0.01* |

Abbreviation: OR, odds ratio; CI, confidence interval;

## Discussion

This study shows that transthyretin amyloidosis cardiomyopathy (ATTR-CM) appears to be associated with a lower risk of Alzheimer’s disease (AD). This observation persists even after adjustment for major cardiovascular and metabolic risk factors, associated with the risk of dementia.

Several studies have highlighted co-occurrences of amyloid deposits in the heart and brain. For instance, the Hisayama study, a Japanese population-based autopsy cohort, found a correlation between cardiac ATTR deposits and AD pathology in older adults [16]. Similarly, a case report has described dual amyloidosis involving wild-type ATTR-CM and AD diagnosed in vivo in a patient [17]. Very recently, Sugita and collaborators described systemic ATTR in 6 of 50 autopsy cases with AD and cardiac hypertrophy [11]. However, these associations and co-occurences may simply reflect shared age-related pathological processes rather than direct causality.

ATTR-CM and AD both involve protein misfolding and aggregation, with pathophysiological overlap at key stages of amyloidogenesis: imbalance between protein synthesis and degradation, loss of conformational stability, toxic oligomer formation, and failure of clearance systems such as proteasomes, autophagy, and innate immune phagocytes [7,8]. Despite their differences — Aβ is derived from the enzymatic cleavage of amyloid precursor protein, whereas TTR is a tetrameric transporter of thyroxine and retinol, whose dissociation into unstable monomers initiates the pathogenic amyloid process — the aggregation cascade shares similar molecular checkpoints. Furthermore, aging is a critical upstream factor common to both diseases, contributing to proteostasis network exhaustion, impaired protein quality control, and increased oxidative stress, thereby fostering systemic amyloidogenic environments [3,4]. Considering the overlapping pathological framework, the inverse association observed in our study suggests that alternative mechanisms may be at work.

One hypothesis is that central nervous system (CNS) derived TTR retains its neuroprotective properties, even in patients with peripheral ATTR-CM. TTR is synthesized by the choroid plexus and secreted into cerebrospinal fluid, where it can bind Aβ peptides, inhibit fibrillogenesis, facilitate clearance via LRP1, and preserve blood-brain barrier integrity [8,18,19]. In transgenic models, TTR overexpression reduces Aβ burden, tau hyperphosphorylation, and cognitive decline [20]. As hepatic TTR and CNS TTR are compartmentalized by the blood-brain barrier, it is plausible that even in amyloidogenic peripheral states, CNS TTR remains stable and functional [7]. A speculative hypothesis is that ATTR-CM may trigger compensatory upregulation of TTR production in the CNS, potentially increasing CSF concentrations and enhancing Aβ clearance. This notion is supported by findings showing tissue specificities of amyloidosis. Indeed, secretion efficiency differences based on the protein chaperone in different tissues may influence the tissue specificity of extracellular amyloidopathies [21]. Another potential modulator is tafamidis, a TTR stabilizer that is used to treat ATTR-CM. Although it was not evaluated in our study, tafamidis may exert indirect CNS effects by improving systemic proteostasis or reducing circulating misfolded TTR. However, to date, no clinical evidence has confirmed a protective cognitive effect of tafamidis.

Another hypothesis that could explain this negative association between AD and ATTR-CM could be a “diagnostic bias”. Patients with AD may be underdiagnosed for ATTR-CM due to reduced access to cardiologists and in-depth evaluations, cognitive barriers to complex diagnostic procedures, and competing diagnostic priorities [22],[23]. Conversely, early cognitive symptoms in older patients with ATTR-CM may be misattributed to cardiovascular deconditioning and not investigated further. To address part of these biases, we looked at whether the number of hospitalizations – which provide diagnosis ICD-10 codes, differed between cases and controls and between patients with and without AD and models were adjusted for the number of hospitalizations. If this analysis controlled for hospitalization rates aimed to minimize this bias, it cannot be entirely excluded. Furthermore, patients with AD may potentially be undertreated with tafamidis.

Our study has several strengths. One of the major strengths is the large and well-characterized cohort of patients with ATTR-CM, probably one of the largest samples of ATTR-CM patients with more than 19,000 patients. We relied on a robust identification algorithm developed by Damy et al [13] to accurately capture cases within the French National Health Data System (SNDS), ensuring diagnostic specificity and epidemiological relevance. Moreover, the prevalence of AD in our control population was 5.0%, which is globally consistent with French epidemiological data on dementia among individuals over 65 years of age. Specifically, prior studies report an overall AD prevalence of approximately 6.1% in men and 8.9% in women in this age group [24,25]. Direct comparison with these other studies should be considered with caution, firstly because prevalence was measured over five years and across a wide age range, and secondly because the characteristics of our control population are biased in relation to the general population, as they are matched to the cases. Indeed, males represent 70% of the sample, due to higher prevalence of ATTR-CM in males [26], in which prevalence of AD is lower [27]. However, this prevalence of AD in controls globally supports the validity of our control group and strengthens the interpretation of the lower prevalence in the ATTR group. Another key strength is the rigorous matching strategy implemented in the design. Cases and controls were matched on crucial demographic and clinical variables, including age, sex, hypertension status, and geographical region, minimizing confounding and enhancing comparability between groups. We also matched with the risk set sampling method accounting for high competing risks of mortality in this specific population. Extensive multivariable adjustments for known dementia risk factors were also conducted, in alignment with the comprehensive framework proposed by Livingston and collaborators [15]. This includes cardiovascular, metabolic, sensory, and psychiatric comorbidities, allowing us to isolate the specific association between ATTR-CM and AD more precisely.

Despite its strengths, our study presents several limitations that must be acknowledged. First, the observational and cross-sectional design precludes any inference of causality. While we identified a significant association between ATTR-CM and a lower prevalence of AD, we cannot determine the directionality of this relationship, nor exclude residual confounding. Another important limitation is the absence of clear consensual definition of AD, and notably biomarker-based definition. Our diagnoses relied on administrative codes within the SNDS database, based on ICD-10 codes, which do not capture biomarker-confirmed AD. This may introduce misclassification, particularly in older adults whom cognitive impairment can have multiple origins. These limitations underscore the need for prospective, biomarker-based studies to further validate and explore the relationship between ATTR-CM and AD. Some comorbidities included as covariates are clearly under-estimated based on ICD-10 codes (e.g. alcohol use or tobacco smoking), but this underestimation is probably the same in both groups (case and controls). Then, to limit misclassification bias inherent to ICD-10 coding in the SNDS, we elected to systematically exclude all patients carrying a code for CAA. The algorithm used to identify cardiac amyloidosis (ICD-10 E85 combined with cardiovascular codes and/or tafamidis dispensing) remains exposed to a nosological overlap between “cardiac amyloidosis” and “cerebral amyloidosis,” creating potential coding errors. Including these records would have artificially inflated the ATTR-CM group characterized by a markedly higher prevalence of Alzheimer’s disease. Our decision therefore prioritizes the **s**pecificity of the ATTR-CM phenotype to preserve the validity of the exposure, at the cost of a limited risk of excluding a small number of true ATTR-CM/CAA co-occurrences. We state this restriction as a study limitation, emphasizing that it reflects an administrative coding context and aims to reduce differential misclassification that could affect the primary endpoint. Sensitivity analysis conducted without exclusion of CAA in tafamidis-treated cases and controls groups increased the strength of the association but not significantly.

Future research is needed, notably mechanistic studies exploring cardiac and cerebral amyloidogenesis. Animal models, in-vivo human studies with biofluid and imaging biomarkers, and post-mortem studies, could help elucidate the systemic versus compartmentalized regulation of amyloidogenic processes, according to the different proteins and organs involved. Particular attention should be paid to the aging proteostasis network and its differential vulnerability across tissues, to better understand potential anti-amyloid neuroprotective roles of TTR and the absence of bi-compartmental protein aggregation.

## Conclusion

This nationwide population-based case-control study in a very large sample of ATTR-CM patients reveals a significant inverse association between ATTR-CM and AD. These findings may support the hypothesis of a potential pathophysiological interaction of ATTR-CM and AD, notably at the protein level from the protein folding to the formation of insoluble amyloid fibrils. However, diagnostic or prescription-related biases cannot be ruled out and warrant caution in interpretation. This first step – at the population level – need to be confirmed, and enhanced by future studies at the molecular, biochemical and clinical levels. Our results highlight the need for further mechanistic and longitudinal research to better understand the potential interactions between systemic amyloidosis, brain amyloidosis and neurodegeneration. Identifying shared pathways or therapeutic targets could pave the way for innovative strategies in managing these two aging-related diseases.

## Data Availability

The data used in the study cannot be shared publicly because the French law on access to individual health data. Access to National Health database (SNDS) data requires to follow a process, the steps of which are described on the website of the French Health Data Hub (https://www.health-data-hub.fr/).

## Author Contributions

Study concept and design: Bretelle, Gervais, Dauphinot, Garnier-Crussard.

Acquisition, analysis, or interpretation of data: All authors.

Drafting of the manuscript: Bretelle, Gervais, Garnier-Crussard

Critical revision of the manuscript for important intellectual content: All authors.

Statistical analysis: Gervais

Study supervision: Garnier-Crussard.

## Disclosure statement

Independent of this work, Fannie Bretelle is an unpaid sub-investigator for clinical trials sponsored by Novo Nordisk, Medesis Pharma and GlaxoSmithKline. She declares that he has not received any personal funding and has not participated in any remunerated activities. Michel Chuzeville received travel grant from Astra Zeneca, and expert fees from Pfizer, Novartis, Lilly, Astra Zeneca, Bristol Myers Squibb. Antoine Garnier-Crussard is an unpaid sub-investigator or local principal investigator for clinical trials and studies sponsored by UCB Pharma, Biogen, TauRx Therapeutics, Roche, Novo Nordisk, Alzheon, Medesis Pharma, GlaxoSmithKline, and received research grant from Roche, Lilly and Eisai. He declares that he has not received any personal funding and has not participated in any remunerated activities.

Other authors have no disclosure to report.

## Funding

No specific fundings. The authors of this manuscript are permanent or temporary employees of the Hospices Civils de Lyon and/or the University of Lyon 1.

## Notes

### Funding Statement

No funding

### Author Declarations

This study is a population-based nested case-control study within the electronic healthcare French database (Systeme National des Donnees de Sante, SNDS). SNDS is a real-world database which covers 99% of the French population. Because the study involved neither direct experimentation on humans nor the use of human biological samples, and relied solely on pre-existing anonymized administrative data, neither written informed consent from participants nor authorization from an institutional ethics committee was required under of the French Public Health Code.

