## Supplementary Material for "Cardiac amyloidosis and the risk of Alzheimer’s disease: a population-based nested case-control study"

- **eTable 1. Number of hospitalizations in AD Patients and Non-AD patients**
- **eTable 2. Full adjusted multivariate model**
- **eTable 3. Sensitivity analysis with the inclusion of patients with cerebral amyloid angiopathy**

**eTable 1. Number of hospitalizations in AD Patients and Non-AD patients**

| **Characteristic** | **Study Participants, No. (%)** | |  |
| --- | --- | --- | --- |
|  | **AD Patients**  **(n = 4 436)** | **Non-AD**  **(n = 91 764)** | ***p-value*** |
| Hospitalizations, mean (SD)^a^ | 5.8 (7.4) | 5.5 (22.5) | *0.24* |
| Abbreviation: AD, Alzheimer’s disease  ^a^ Mean number of hospitalisations in the 5 years before index date | | |  |

**eTable 2. Full adjusted multivariate model**

| **Variable** | | **All participants** | |
| --- | --- | --- | --- |
|  |  | **Adjusted OR (99% CI)** | **p-value** |
| Alzheimer's disease | | | |
|  | Yes | 0.65 (0.56-0.75) | <0.01 |
| Atrial fibrillation | | | |
|  | Yes | 4.21 (3.97-4.46) | <0.01 |
| Stroke | | | |
|  | Yes | 1.32 (1.17-1.49) | <0.01 |
| Heart failure | | | |
|  | Yes | 8.20 (7.70-8.73) | <0.01 |
| Diabetes | |  |  |
|  | Yes | 0.88 (0.83-0.94) | <0.01 |
| Dyslipidemia | |  |  |
|  | Yes | 1.54 (1.44-1.64) | <0.01 |
| Obesity | |  |  |
|  | Yes | 0.86 (0.79-0.93) | <0.01 |
| Hearing loss | |  |  |
|  | Yes | 1.15 (0.98-1.35) | 0.03 |
| Depression | |  |  |
|  | Yes | 0.82 (0.77-0.)88 | <0.01 |
| Smoking | |  |  |
|  | Yes | 0.71 (0.52-0.97) | <0.01 |
| Alcohol use | |  |  |
|  | Yes | 0.88 (0.63-1.24) | 0.35 |
| Glaucoma | |  |  |
|  | Yes | 1.13 (1.05-1.21) | <0.01 |
| Aged-related Macular Degeneration | | | |
|  | Yes | 0.90 (0.80-1.01) | 0.02 |
| Hospital stays (1 unit rise) | | |  |
|  | 0 | 1 [Reference] |  |
|  | 1 | 1.00 (0.99-1.01) | 0.55 |

For each variable “No” is the reference.

Abbreviation: OR, odds ratio; CI, confidence interval;

**eTable 3. Sensitivity analysis with the inclusion of patients with cerebral amyloid angiopathy**

| **Variable** | | **All participants** | |
| --- | --- | --- | --- |
|  |  | **Adjusted OR (99% CI)** | **p-value** |
| Alzheimer's disease | | | |
|  | Yes | 0.56 (0.49-0.65) | <0.01 |
| Atrial fibrillation | | | |
|  | Yes | 3.10 (2.92-3.28) | <0.01 |
| Stroke | | | |
|  | Yes | 1.31 (1.19-1.43) | <0.01 |
| Heart failure | | | |
|  | Yes | 8.67 (8.15-9.22) | <0.01 |
| Cerebral amyloid angiopathy | |  |  |
|  | Yes | 1.85 (1.06-3.25) | <0.01 |
| Diabetes | |  |  |
|  | Yes | 1.22 (1.15-1.29) | <0.01 |
| Dyslipidemia | |  |  |
|  | Yes | 1.41 (1.32-1.50) | <0.01 |
| Obesity | |  |  |
|  | Yes | 0.79 (0.73-0.86) | <0.01 |
| Hearing loss | |  |  |
|  | Yes | 1.31 (1.15-1.50) | <0.01 |
| Depression | |  |  |
|  | Yes | 0.95 (0.89-1.01) | 0.02 |
| Smoking | |  |  |
|  | Yes | 0.83 (0.62-1.10) | 0.08 |
| Alcohol use | |  |  |
|  | Yes | 0.98 (0.72-1.33) | 0.83 |
| Glaucoma | |  |  |
|  | Yes | 1.10 (1.03-1.18) | <0.01 |
| Aged-related Macular Degeneration | | | |
|  | Yes | 0.91 (0.82-1.02) | 0.04 |
| Hospital stays (1 unit rise) | | |  |
|  | 0 | 1 [Reference] |  |
|  | 1 | 0.99 (0.98-1.01) | 0.03 |

For each variable “No” is the reference.

Abbreviation: OR, odds ratio; CI, confidence interval;

In this sensitivity analysis, we kept patients with cerebral amyloid angiopathy both in cases (in the tafamidis group but not in the non-tafamidis group) and controls. CAA represented 19,232 patients coded in our total population aged 65 years and over.
